# Genomic stratification of clozapine prescription patterns using schizophrenia polygenic scores

**DOI:** 10.1101/2022.02.18.22271204

**Authors:** Djenifer B. Kappel, Sophie E. Legge, Leon Hubbard, Isabella R. Willcocks, Adrian King, John Jansen, Marinka Helthuis, Michael J. Owen, Michael C. O’Donovan, James T.R. Walters, Antonio F. Pardiñas

## Abstract

**BACKGROUND:** Treatment-resistant schizophrenia (TRS) affects ∼30% of individuals with the disorder. Clozapine is the medication of choice in TRS but optimizing administration and dose titration are complex. The identification of predictive factors that influence clozapine prescription and response, including genetics, is of clinical interest in a precision psychiatry framework. We aimed to determine if a polygenic risk score (PRS) for schizophrenia is associated with the highest drug dose an individual received during clozapine treatment.

**METHODS:** We used generalized linear regression models accounting for demographic, pharmacological, and clinical covariates to determine the relationship between PRS and highest daily dose of clozapine. We used two independent multi-ancestry samples of individuals from the UK from a clozapine monitoring system, CLOZUK2 (N= 3133) and CLOZUK3 (N= 909). Schizophrenia PRS were calculated using the latest available GWAS summary statistics from the Psychiatric Genomics Consortium. In a secondary analysis of the two merged cohorts, logistic regression models were used to estimate the relationship between schizophrenia PRS and clozapine doses classified as low, standard, or high (>600 mg/day).

**RESULTS:** After controlling for relevant available covariates, schizophrenia PRS were correlated with the highest clozapine dose ever prescribed, in both CLOZUK2 (β= 12.217, s.e= 3.776, P= 0.001) and CLOZUK3 (β= 12.730, s.e= 5.987, P= 0.034). In the secondary analysis, the schizophrenia PRS was specifically associated with taking a clozapine dose greater than 600 mg/day (OR= 1.279, P= 0.006).

**CONCLUSIONS:** Schizophrenia PRS is associated with the highest clozapine dose ever prescribed in two independent multi-ancestry samples from the UK, suggesting that the genetic liability to schizophrenia might index factors associated with therapeutic decisions in TRS cohorts.

## INTRODUCTION

Approximately one-third of individuals with schizophrenia experience symptoms that do not meaningfully improve after two courses of standard antipsychotics, a presentation often called treatment-resistant schizophrenia (TRS) (1). Clozapine, a second-generation antipsychotic, is the evidence-based treatment of choice for TRS, given its proven efficacy to treat symptoms that have not responded to other antipsychotics (2). However, despite its clear benefits in TRS, clozapine also has the potential to cause a range of adverse effects, and these are major contributors to treatment discontinuation (3). Blood dyscrasias such as neutropenia and agranulocytosis are particularly important adverse drug reactions (ADRs) due to their unpredictability and potentially fatal consequences if unmanaged, and thus regular hematological monitoring has been implemented to mitigate their risk (4). These and other factors complicate clozapine administration and dose titration, contributing to the fact that many eligible patients are not offered clozapine as a treatment option (5). Moreover, it is estimated that only about 50% of those treated respond to clozapine (6), and few objective predictors of therapeutic response or adverse effects have been identified to date (7).

Individual differences in response to pharmacological treatment are known to be influenced by genetic and environmental factors (8,9). Pharmacogenomic research aims to identify genetic variants that contribute to this variability and is one of the most promising pillars of precision medicine strategies (10). To date, while most known pharmacogenomic variants are associated with ADME processes (absorption, distribution, metabolism, and excretion), markers associated with disease and disorder risk can also be assessed to investigate treatment outcomes (11). In this sense, composite metrics of genetic risk such as polygenic risk scores (PRS), which combine the weighted effects of many variants across the genome, have become widely used in medical genomic research and are also seen as potential predictive markers which could eventually be introduced into patient care (12,13).

Hundreds of schizophrenia susceptibility loci have been identified by large-scale GWAS, and these have pointed to neurobiological pathways and mechanisms likely to be disrupted in the disorder (14). Several of these could feasibly play a role in antipsychotic treatment response, such as the dopaminergic signaling pathways indexed by *DRD2* (15). This suggests that investigating the association between genetic liability to the disorder and response to antipsychotics might be fruitful, with a hypothesis being that heavier genetic burden could be associated with poorer treatment response. As treatment outcomes in schizophrenia are multifaceted and depend on complex factors including treatment adherence (10), many studies on this matter have investigated treatment response by focusing on individuals with the most severe symptomatology, often using samples of those with TRS (16). However, the evidence for schizophrenia risk alleles being predictors of TRS is inconclusive, and the largest study on this topic suggests that the schizophrenia PRS is not substantially elevated in those with this diagnosis (17). However, the possibility remains that genomic liability to schizophrenia could be associated with clozapine treatment outcomes, but this line of enquiry has been hampered by the difficulties of recruiting large samples of those with TRS given the more severe symptomatic burden affecting these individuals, which complicates their involvement in traditional research settings (18).

A key challenge for clinicians is determining the optimal dose of clozapine for a given individual, which requires weighing up the relative likelihoods of therapeutic response versus ADRs. Clinical caution to avoid adverse effects, which even in mild instances can be debilitating, might also lead to individuals spending weeks or months on a given dose without apparent benefits before they are escalated to a higher one (19). Additionally, simply increasing clozapine doses does not warrantee better responses to the drug, with meta-analytic evidence pointing to the need of taking drug metabolism into account in clinical practice (20). In this sense, therapeutic drug monitoring (TDM) schemes, when available, can facilitate fine-tuning clozapine metabolite concentrations (or “levels”) for optimal response, but are particularly suited for the identification of extremely poor or rapid metabolizers (21), a subset of the general population that does not fully account for the rate of clozapine non-responders (22). For these reasons, investigating clinical and demographic characteristics, including genetics, underlying clozapine prescriptions in real-world settings is a pathway towards both the inference of predictive factors for treatment outcomes and a better understanding of the clinical decision-making processes behind clozapine dose escalation, both of which are of clinical interest in a precision psychiatry framework.

The present study analyzes genetic and clozapine pharmacokinetic data in two retrospective cohorts from the CLOZUK project, one of the largest DNA sample collections worldwide of individuals with TRS (23). The aim is to assess whether the schizophrenia PRS, at present a well-powered metric with associations across psychiatric and personality traits (24), is also correlated with the clozapine doses prescribed to those with TRS. Given the underuse of clozapine and the complexities of its clinical management, inferring the potential relevance of genomic information in this setting could be informative for the development of future stratification and drug dosing algorithms. Additionally, novel observations supporting that the schizophrenia genetic liability might also index therapeutic decisions and outcomes would add to a growing body of evidence on the relationship between disease onset, symptomatology and progression; an area of great interest and activity for precision psychiatry research.

## MATERIAL AND METHODS

### SAMPLES

The CLOZUK cohort is comprised of individuals taking clozapine in the UK whose DNA samples were collected anonymously. Collection was facilitated by Leyden Delta B.V (Nijmegen, the Netherlands), one of the providers of clozapine in the UK. For the present research, we accessed data from a wave of data collection termed CLOZUK2, which, unlike an earlier sample (CLOZUK1; 25), was linked to repeated assessments of clozapine pharmacokinetics by the medical laboratory Magna Labs (Ross-on-Wye, England). Additional descriptions of this cohort, genotyping procedures, and data collection have been previously reported (23,26).

A total of 3,439 unrelated individuals over the age of 18 years were available from CLOZUK2 with pharmacokinetic and genotypic data, and more than 12,000 pharmacokinetic assays. This sample was curated to remove individuals taking clozapine for less than 18 weeks in order to ensure steady-state levels of clozapine in plasma had been reached, and to exclude individuals undergoing the initial titration process (19,27). In contrast to previous studies which have focused on European participants to minimize population stratification, we did not filter our data based on self-reported or genetically-inferred ancestry. Our final curated dataset included a total of 3,133 individuals from CLOZUK2. A summary of demographic and clinical characteristics is given in **Supplementary Table 1**.

New to this study, we also report another wave of CLOZUK data (CLOZUK3), with more than 900 individuals and 5,000 pharmacokinetic assays. Its collection follows the procedure detailed earlier for CLOZUK2 (23), including the curation protocol for clozapine levels data (26). For our analyses of CLOZUK3, we did not exclude individuals with a clozapine treatment shorter than 18 weeks, as that information was not available, though we note in CLOZUK2, those individuals represent a minority (9.3%) of the sample. However, to increase compatibility with the curation procedures of CLOZUK2 and reduce the likelihood of analyzing individuals going through clozapine initiation/titration, we removed those with a highest clozapine dose <100 mg/day. Our final CLOZUK3 dataset included genetic and pharmacokinetic data for 909 individuals (**Supplementary Table 1)**.

All procedures contributing to this work comply with the ethical standards of the relevant national and institutional guidelines (UK National Research Ethics Service approval ref. 10/WSE02/15, following UK Human Tissue Act).

### STUDY DESIGN

For the primary analyses, we focused on the highest clozapine daily dose for each of the individuals included in our samples, CLOZUK2 was analyzed first and treated as a discovery sample, while the smaller CLOZUK3, collected afterwards and genotyped separately, was used as a replication sample (**Figure 1**).

**Figure 1.**
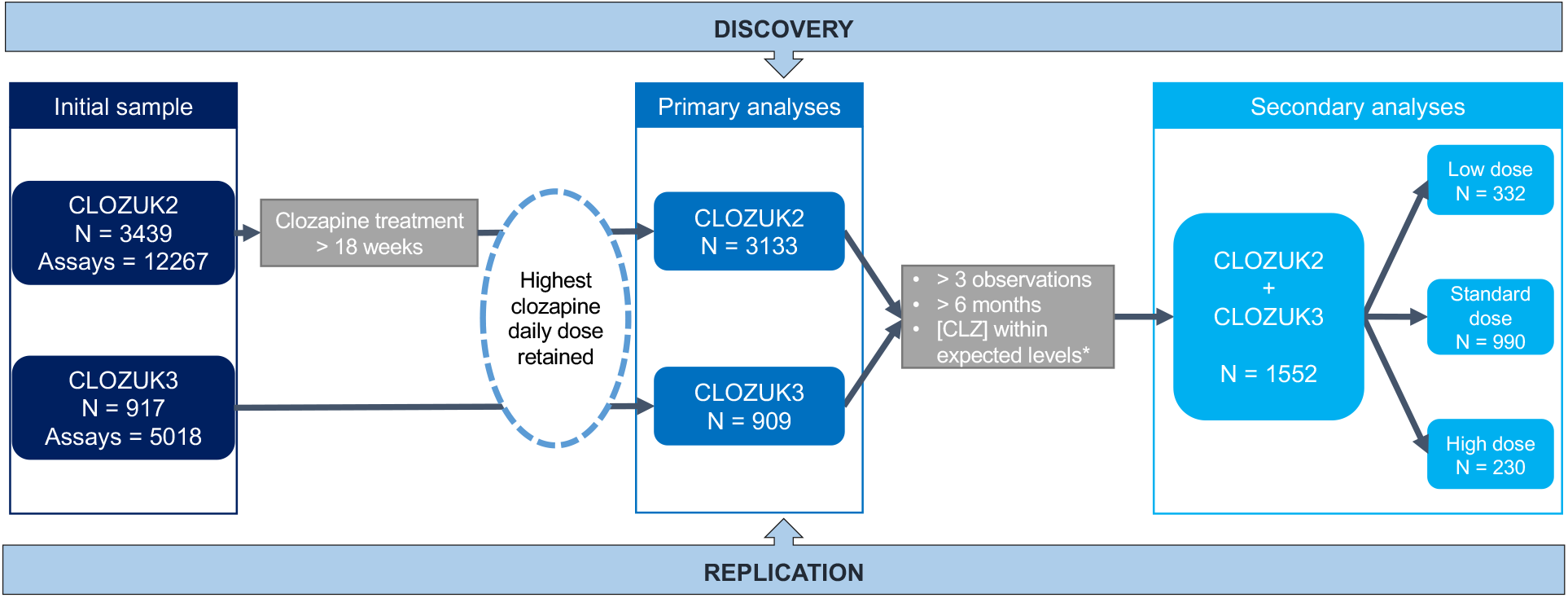
Sample inclusion flowchart. Curation procedures relevant to each analyses step for both CLOZUK2 and CLOZUK3 are represented. [CLZ] = clozapine plasma levels *dose-adjusted expected levels extracted from Couchman et al. 2010.

In secondary analyses, we merged the two CLOZUK cohorts and then stratified these individuals by their highest daily clozapine dose into three categories: (1) those taking a low dose (<300 mg/day), (2) those taking standard maintenance doses (300-600 mg/day), and (3) those individuals taking a higher dose than the usual maintenance dose (>600 mg/day)(28). For these analyses, we also included extra curation procedures (**Figure 1**): First, we selected only those individuals with at least three assays in the clozapine monitoring system, spanning a period of 6 months or more. This step aimed for the clozapine dose we assessed to more accurately reflect the real highest dose a participant was likely to have taken throughout their treatment. Second, we removed individuals likely not taking their medication (non-adherence) and/or presenting an atypical (rapid/poor) clozapine metabolism, as prescription patterns in these individuals would not likely follow the linear trends of the general population. This last procedure was done by excluding all assays in which the observed clozapine plasma concentrations did not match those expected for the recorded clozapine daily dose, as reflected in Table 5 of Couchman et al., 2010 (29).

### GENETICS

The genotyping of CLOZUK2 was conducted using Illumina^®^ HumanOmniExpress arrays. A detailed description of genotyping, quality control and imputation procedures for genomic data can be found elsewhere (23). The CLOZUK3 cohort was genotyped using the Illumina® Infinium Global Screening Array-24 and curated using the DRAGON-Data *“GenotypeQCtoHRC”* pipeline (30), based on PLINK v1.9 (31). Briefly, this pipeline includes basic genotypic quality control (QC) allowing for 5% of missing individual and marker data (32), checks for sample identifiability including the sex chromosomes (33), and assessment of genotyping errors by removal of markers outside Hardy-Weinberg equilibrium (mid p >10^−6^; (34)). Imputation of CLOZUK3 was performed on the Michigan Imputation Server with the HRC reference panel and default options (35). For PRS analyses, imputed CLOZUK2+3 dosages were converted to best-guess genotype calls (Genotype probability > 90%; INFO > 0.9, minor allele frequency [MAF] > 10%, HWE mid p > 10^−4^). For the merging of CLOZUK2+3 genomic data, we also followed the best practices implemented in *“GenotypeQCtoHRC”*, including the removal of all strand-ambiguous or non-overlapping SNPs and the recalculation of MAF thresholds in the whole sample.

For deriving the main predictor of interest, we computed genome-wide PRS profiles from the latest schizophrenia multi-ancestry meta-analysis from the PGC, which has a majority of European participants (14). Given the CLOZUK2 cohort was included in this publication, to avoid sample overlap between training and testing sets, we derived deduplicated schizophrenia summary statistics before calculating PRS using the PGC analytic infrastructure. However, we used summary statistics from the full PGC GWAS as training set to derive PRS in CLOZUK3 as that sample was not included in the PGC meta-analysis. As data within CLOZUK does not include known proxies of demographic and lifestyle factors associated with drug metabolism (36), we extracted summary statistics for coffee intake, body mass index (BMI), and smoking behavior from the GIANT consortium (37) and the GeneATLAS UK Biobank GWAS resource (38). All summary statistics were restricted to SNPs with MAF>10% and INFO>0.9, matching post-imputation genotype curation. PRS were computed using the PRScs method (39), adjusted for LD-structure with default options and a shrinkage parameter of phi=1 for schizophrenia (24) and phi=auto otherwise. Before statistical analysis, each PRS was standardized (mean = 0, SD = 1) to facilitate the interpretability of the results.

### STATISTICS

#### Primary Analyses

To analyze the association between the schizophrenia PRS and the highest daily dose of clozapine, we used generalized linear regression models accounting for relevant demographic, pharmacological, and treatment covariates. In our main analysis, these included sex, age and age^2^, all present in the CLOZUK records, and PRS metrics as proxies of BMI, coffee intake and smoking habits. In further analyses, we also included two other predictors which might affect the outcome of highest dose, and which could potentially act as mediators of our observed effects. The first predictor was the clozapine plasma concentrations observed at the point of highest dose. A moderating effect of these concentrations would be consistent with schizophrenia risk variants reflecting features of drug metabolism, a hypothesis which has been explored in previous research on TRS (40). The second predictor was the number of times each individual’s clozapine levels were assessed in the complete dataset. This could be a proxy for potential confounders as, in order to monitor adherence and anticipate level-dependent ADRs, pharmacokinetic monitoring is likely to be requested more frequently as individuals’ clozapine doses escalate, thus increasing the likelihood of those on high doses being present in our dataset.

All regression analyses were conducted in the statistical software R v4.1.0. The change in R^2^ due to the inclusion of each covariate (also known as semi-partial R^2^ or ΔR^2^) was estimated as an index of the proportion of variance explained by any individual factor in our model using the “rockchalk” package (41).

All CLOZUK2 individuals underwent a previous genetic ancestry analysis using Ancestry Informative Markers (AIMs) and a Linear Discriminant Analysis (LDA) model (42), which we reproduced in CLOZUK3. From this procedure we derived, for each individual, LDA-based probabilities of belonging to five biogeographic regions (Europe, East Asia, Southwest Asia, North Africa and Sub-Saharan Africa). To account for potential confounding from population stratification, we included these probabilities in all our regression models, plus the first ten principal components (PCs) from a PC-AiR analysis (43).

#### Secondary Analyses

We also undertook a series of analyses focusing on a broad but clinically relevant categorization of clozapine dose. We used multinomial and binary logistic regression models to estimate the effects of the schizophrenia PRS in the probability of taking highest clozapine doses within three different dose groups: low (<300 mg/day), standard (300-600 mg/day) and high (>600 mg/day) (28). We fit three separate pairwise regression models to assess differences between groups, using the same covariates in these models than in the primary analyses. Also, to ensure compatibility between the CLOZUK2 and CLOZUK3 PRS and PCA variables, we used the deduplicated PGC summary statistics as the PRS training set in this secondary analysis and generated the scores and principal components on strictly overlapping markers passing all quality-control filters in the merged sample. As a sensitivity analysis, we also collapsed individuals taking doses in the low and standard range into one category and compared them with those taking high doses (>600 mg/day), as prescribing a high clozapine daily dose generally requires more complex clinical considerations given the likelihood of ADRs than switches within lower thresholds. In this model, we calculated the area under the curve (AUC) from receiver operating characteristic (ROC) curves using the pROC package (44) in R. This is as a rough estimate of the added utility of our genetic predictor when combined with standard demographic variables used in clinical prediction modelling (45,46). We also compared this AUC with the analogous estimate from a model including only demographics (i.e., age, age^2^, sex, and self-reported ethnicity), and another model including demographics and clozapine plasma concentrations to quantify the increase in predictive ability provided by genetic information alone. All the AUC values were calculated exclusively on the CLOZUK2 individuals to take advantage of the self-reported ethnicity information only available for this sample. This has been considered an important predictor for clozapine dose optimization (47) and is not always directly comparable to genetic ancestry estimates.

## RESULTS

### Primary Analysis: Association of the schizophrenia PRS and the highest clozapine dose

We observed a positive correlation between the schizophrenia PRS and highest clozapine dose in CLOZUK2 (β= 12.217, [95% CI: 4.816–19.618], P= 0.001), where the variance explained by the schizophrenia PRS was Δ*R*^2^ ∼ 0.32%. Effect sizes expressed as the change in clozapine dose (mg/day) for one unit increase of the main predictors, accounting for other model covariates, can be seen in **Figure 2A**. These results were essentially unchanged when accounting for possible mediators such as clozapine plasma concentrations and the frequency of clozapine monitoring in our dataset (**Supplementary Table 2**). Interestingly, although there was not an obvious linear effect between the monitoring frequency and the schizophrenia PRS, we did observe larger clozapine daily doses in individuals who underwent monitoring more frequently (**Supplementary Figure 1**), consistent with our initial expectations.

**Figure 2.**
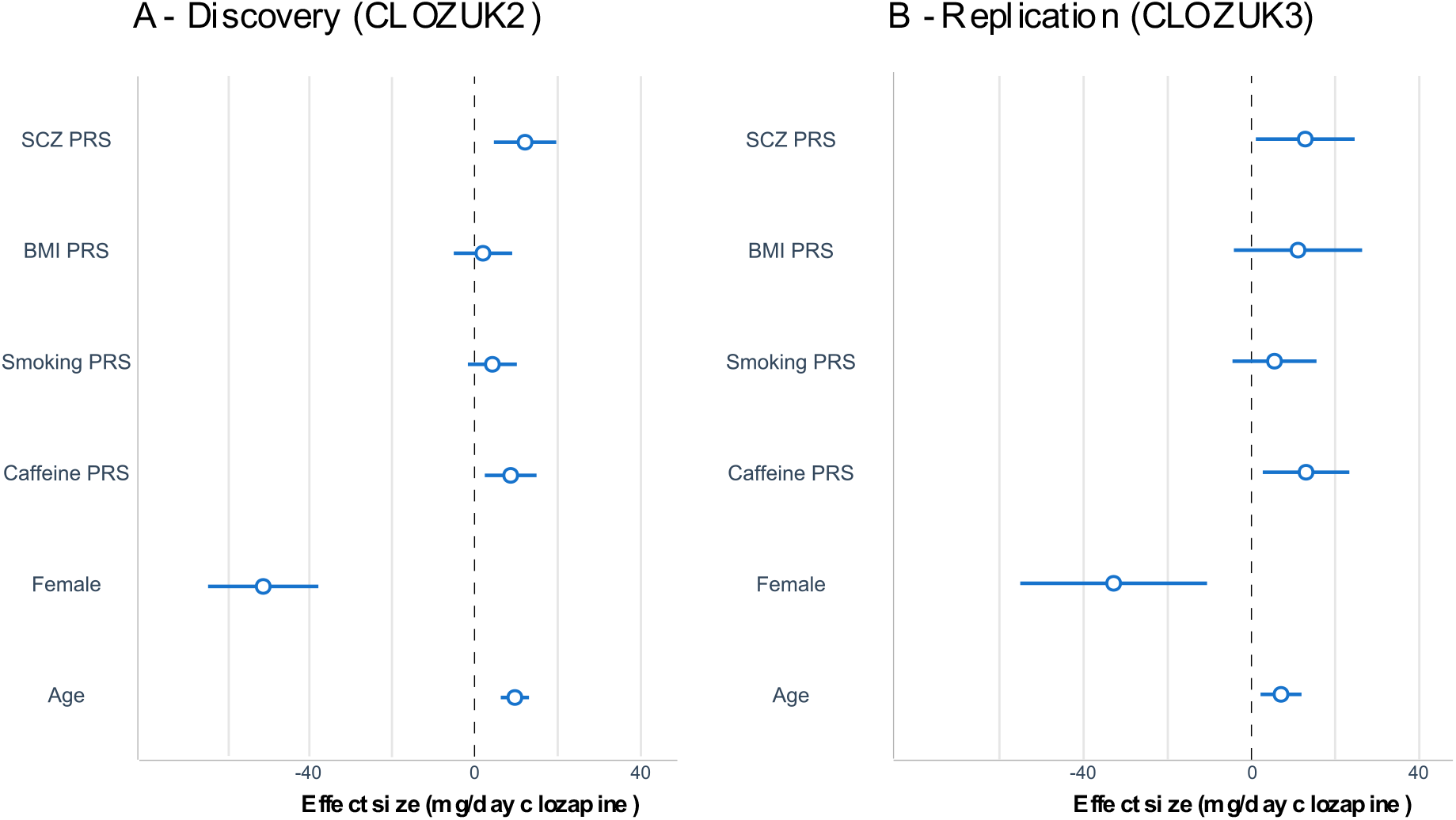
Effect estimates for the predictors of highest clozapine daily dose. Mean coefficient estimates (±95% confidence intervals) are plotted for (A) CLOZUK2, and (B) CLOZUK3.

Exploring our results further via sensitivity analyses, we saw little change in our schizophrenia PRS association by restricting the sample to only individuals of European genetic ancestry (n=2577; β= 11.46, [95% CI: 3.169-19.75], P = 0.007), and established that this result is specific to schizophrenia genetic liability by assessing a wider range of psychiatric, cognitive and personality PRS, none of which were significantly associated with clozapine doses (**Supplementary Figure 2**). We also confirmed that our main detected effects were still consistent and significant when restricting our dataset to observations at clozapine plasma concentration thresholds investigated in previous research on therapeutic response to the drug (20), either between 350-600 ng/L (n= 1089; β= 14.952, [95% CI: 3.167-26.737], P= 0.0131) or above (n= 1283; β= 17.258, [95% CI: 5.823-28.692], P= 0.0031).

To validate our findings in an independent dataset, we replicated this analysis in CLOZUK3 and found results of similar magnitude and sign (β= 12.730, [95% CI: 0.996-24.464], P= 0.033, Δ*R*^2^ ∼ 0.48%; **Figure 2B**), even after controlling for possible mediators (**Supplementary Table 2**).

### Secondary analysis: Genetics-informed classification model of clozapine doses

We next explored to what extent the schizophrenia PRS could reflect broad clozapine prescription patterns in the complete CLOZUK cohort by using a multinomial regression model similar to standard classification algorithms (**Figure 3**). For this, in stratified analyses by clozapine dose categories, we observed that the schizophrenia PRS was associated with the probability of taking high doses when compared to those taking either standard doses (OR = 1.277, 95% CI [1.066-1.530], P = 0.008) or low doses (OR = 1.280, 95% CI [1.029 -1.593], P = 0.027).

**Figure 3:**
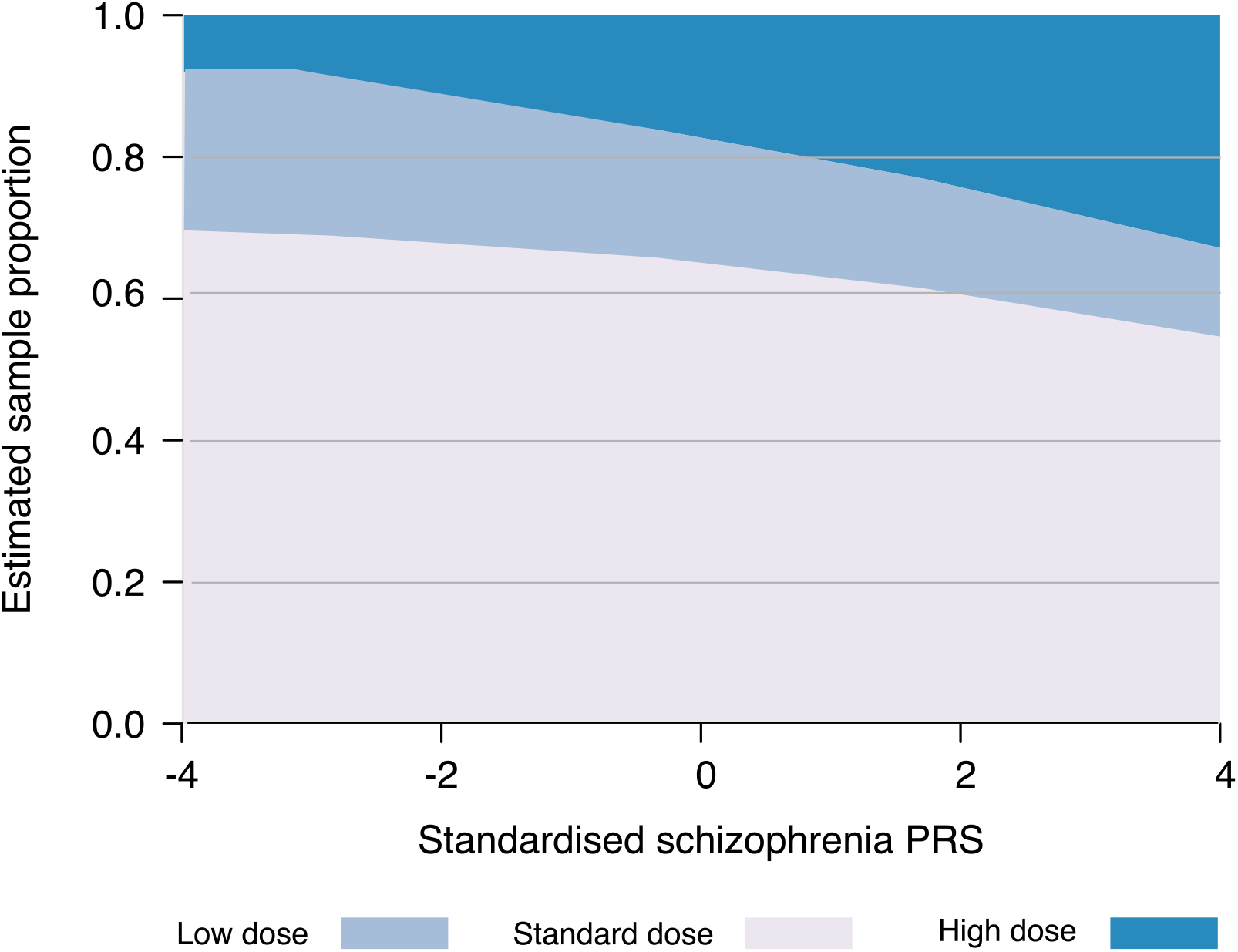
Probability estimates for each of highest clozapine dose categories according to schizophrenia PRS. Y-axis represents the probability of pertaining to each of the 3 doses groups and their estimates along the PRS spectrum.

A second stratified analysis specifically examined differences in those taking clozapine doses over 600 mg/day against those below this threshold. In this analysis, we observed an association between the schizophrenia PRS and probability of taking high doses (OR= 1.279, 95% CI [1.076-1.522], P= 0.005). These results are shown in **Table 1** and as a logit probability curve in **Figure 4**. As an illustration of the detected effects, while the overall prevalence of individuals taking a high dose of clozapine was 15% in the entire CLOZUK sample, it surpassed 20% on those above 2 standard deviations of the schizophrenia PRS, reaching 30% at the upper end of the PRS distribution.

**Table 1:**
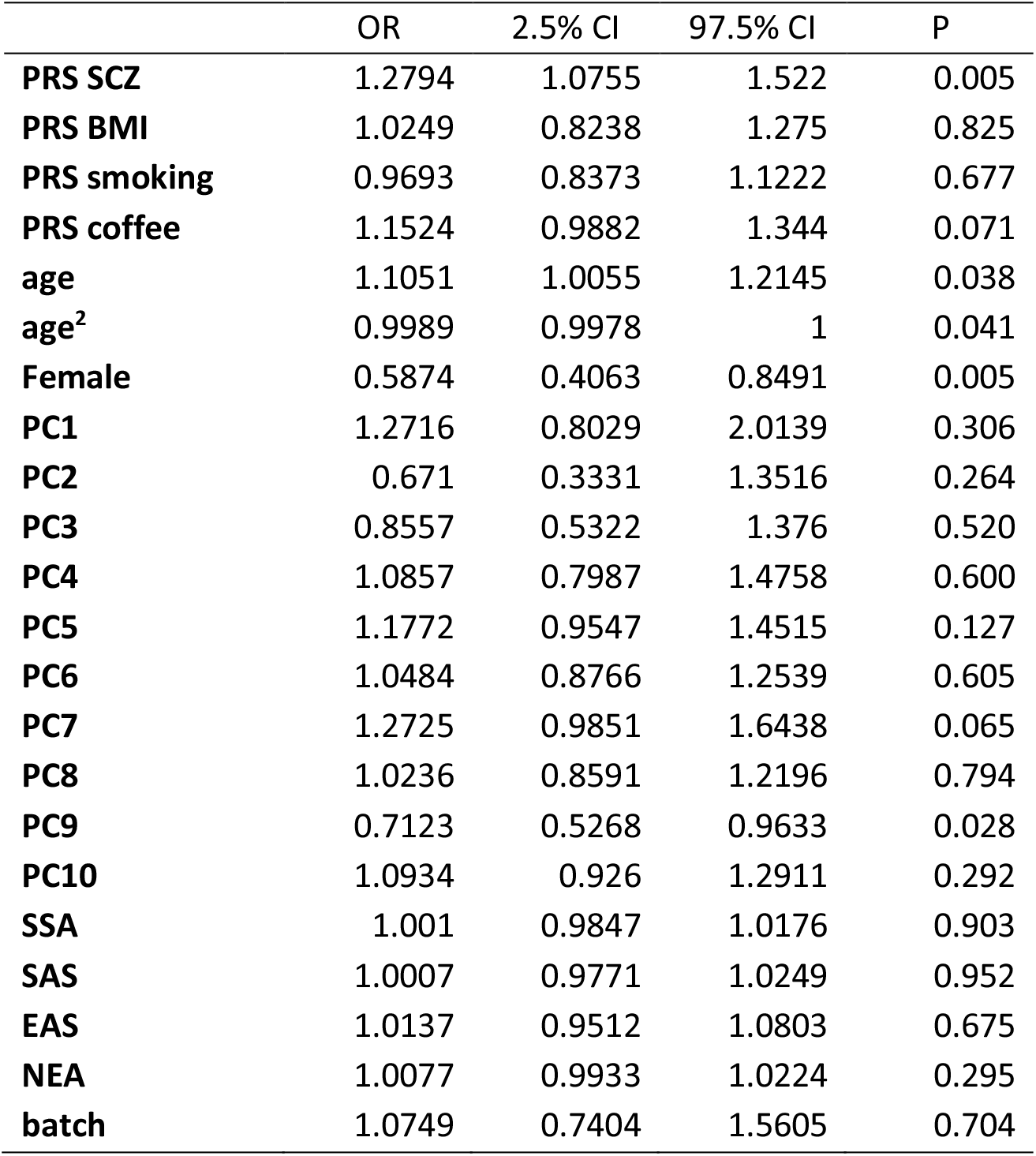
Effect sizes of each predictor included in the model in relation to probability of taking a high clozapine dose.

**Figure 4.**
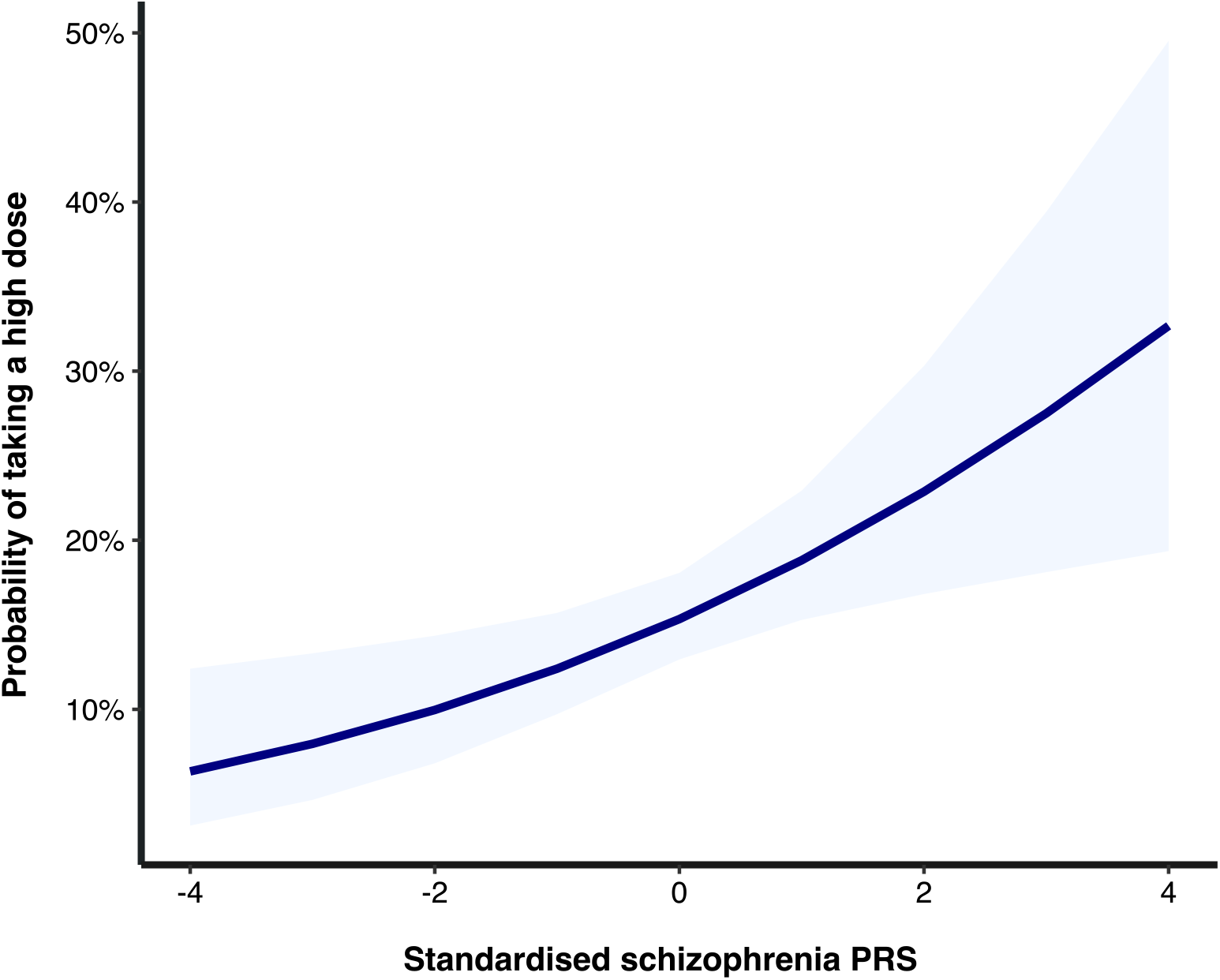
Odds ratios for taking a high clozapine daily dose at different levels of schizophrenia PRS (banded area represents ±95% confidence intervals).

Finally, we assessed the sensitivity and specificity of our prediction models for high clozapine doses by calculating the AUC from ROC statistics, presented in **Supplementary Figure 3**. AUC from the model including all covariates in the previous analysis (**Figure 4, Table 1**) was 0.64, while the demographics-only model (not including any genetically derived covariate) was 0.58. We also show that even when accounting for clozapine plasma concentrations, a known target of dose optimization for clozapine and a strong correlate of actual doses, the inclusion of genetic information marginally improves prediction accuracy.

## DISCUSSION

This study examines whether the polygenic risk for schizophrenia is associated with the daily dose of clozapine in two independent multi-ethnic TRS cohorts. Our main result demonstrates association between genetic liability to schizophrenia, indexed by its PRS, and clozapine highest dose. To our knowledge, this is the first study to report this association. A previous study, albeit using a limited sample (N=44 individuals) did not find an association with schizophrenia PRS and clozapine dose (27). In the secondary analysis, schizophrenia PRS was associated with the probability of taking a high clozapine dose (>600 mg/day). Individuals with high genetic risk had a two-fold increased probability of taking high doses as those on the lower end of the schizophrenia PRS spectrum **(Figure 3)**.

Identifying individuals who are more or less likely to respond to different pharmacological treatments has long been one of the hoped for applications of PRS in precision medicine (12,46). In this study, we show that individuals in the high end of the schizophrenia PRS spectrum are more likely to be prescribed higher clozapine doses than the usual maintenance thresholds (300-600 mg/day). This implies, in the first instance, they have could have been considered to need a higher dose to obtain a therapeutic response. This interpretation is supported by the findings of Frank et al. (2015), who reported that the highest schizophrenia PRSs in a TRS cohort occurred in individuals who are non-responders to clozapine. However, alternative explanations are also possible, such as the schizophrenia PRS delimiting a subset of individuals with greater tolerance to clozapine or simply more prone to either accepting or proposing a dose escalation. While these are arguably speculative, in any case, it is unlikely that genetic make-up is the only or even a major contributor in the very complex network of factors that impact antipsychotic response. This warrants caution in the interpretation of our findings’ potential clinical relevance, which needs to be evaluated further once larger datasets become available.

The mechanisms by which schizophrenia genetic liability may affect response to clozapine treatment and real-world prescription patterns are unknown. It is plausible that part of the common genetic variance linked to schizophrenia might affect overlapping pathways involved in pharmacodynamics (40), the genomics of which are relatively unknown in psychopharmacology (48). It is also known that some of the genes identified in schizophrenia GWAS are related to neurodevelopment, synaptic biology, and immune response, pathways that have been linked to antipsychotic response and adverse effects (49,50). Given that studies suggest the efficacy of clozapine could be related to targeting TRS-specific biological pathways (16), an interesting perspective would be to analyze if future GWAS focused on TRS or antipsychotic treatment response are themselves associated with clozapine dose.

In this study, we leveraged the longitudinal aspect of CLOZUK and examined the highest clozapine daily dose recorded for each individual during clozapine drug monitoring. We hypothesized that, given official prescription guidelines suggesting that once in the therapeutic dose range, doses should only be increased in case of non-response to the treatment, it is reasonable to postulate that higher dosing could be a proxy phenotype for poorer treatment response in the absence of adverse effects. Although this hypothesis cannot be formally tested in our data, previous studies have suggested that individuals requiring higher doses of clozapine could be an indicator of poor treatment response (20). In any case, our results show that individuals with a high schizophrenia PRS are more likely to be prescribed a high clozapine dose. The a priori knowledge of this information could help guide the titration process to benefit treated individuals more quickly while managing their symptoms. Alternatively, it should be considered that at least some of these individuals might be clozapine non-responders, also known as ultra treatment-resistant, which might inform protocols to assess early whether this drug is likely to be effective for them at all.

The current study’s findings suggest that although we observe a statistically significant association, the variance explained by the schizophrenia PRS is small, and other genomic and non-genomic factors potentially contribute to a larger extent to the final phenotype. We used several sources of genetic information in our models (i.e., PCs, genetic ancestry, and PRS for more than one condition), which combined help explain a non-trivial amount of variance in the highest clozapine dose. In line with this is the notion that PRS alone will likely have a relatively small impact in driving clinical practice even once its practical implementation becomes feasible (12); however, other areas such as cardiovascular disease have shown that combining genetic information with non-genetic predictors and risk factors could be clinically meaningful and may help guide treatment choices (51).

## Strengths and limitations

The strengths of our present study include its large sample size, being one of the largest TRS cohorts in the world with genetic and longitudinal pharmacokinetic information. Another distinctive feature was our independent replication in another sample with the same collection protocol and characteristics. Additionally, we used all the individuals in the CLOZUK cohort with available data, regardless of ancestry, expanding our analyses to non-European populations traditionally underrepresented in genomics research.

Several limitations of the current study also need to be considered, and results should be interpreted in light of those. First, this sample is based on electronic health records collected during mandatory clozapine monitoring, which do not include contextual information on clinical management, treatment response, or lifestyle. This impacted our ability to determine factors known to influence clozapine metabolism in our assessed individuals, including smoking status, weight, regular caffeine use, or the use of other medications. However, we attempted to mitigate these issues by using PRS to derive genetically informed proxies of these as in a previous study (26), and these indeed contributed to explain part of the variance in our dataset (**Figure 1, Supplementary Table 2, Supplementary Table 3**). Second, as in all retrospective analyses, unmeasured confounders might have an influence on the effects detected, although all models were adjusted for known potential confounders in primary and sensitivity analyses. Third, we acknowledge that even though we present evidence for an association of schizophrenia PRS with clozapine dose in two independent samples, further research will require additional data on treatment response to evaluate mechanisms linking our observations to real-world prescribing practices. Of this matter it should be noted that even though we included some well-known contributors to the inter-individual variability in clozapine metabolism and dosing, our best full model still only explains 14% of the variance in this phenotype. This suggests most of the factors contributing to clozapine dosing are still elusive. For example, our genetic measures only capture the effects of common genetic variants. The impact of rare variants in genes involved in clozapine metabolism has not been evaluated yet to our knowledge and could account for more of the variability related to drug response (52).

As a final consideration, our main predictor is a schizophrenia PRS built from a mostly cross-sectional case-control analysis, which is not necessarily representative of the diversity of individuals and symptom profiles encompassed by real-world samples of those with schizophrenia or TRS. Also, although our sample is multi-ancestry by design and ascertained through a population-level clozapine monitoring system, it is still primarily composed by European individuals and mainly European GWAS have been used in generating its PRS. For these reasons, it is difficult to evaluate whether potential downstream applications of our research would be translatable to real-world clinical settings or broadly applicable to individuals from worldwide ethnic and genetic backgrounds.

In conclusion, we report that the schizophrenia PRS is associated with highest clozapine dose in patients with TRS in two independent multi-ancestry cohorts, suggesting that genetic susceptibility to schizophrenia is associated with treatment choices in these samples. Despite the ongoing debate over the clinical utility of PRSs in precision psychiatry, our study adds to the growing body of evidence showing that genomic information can lead to novel answers in topics of interest for clinical care (12,53). More studies are now needed to confirm our findings and to benchmark to which extent this or similar data could lead to future improvements in therapeutic decision-making and in the overall clinical management of TRS.

## Data Availability

The data analyzed in this study is subject to the following licenses/restrictions: To comply with the ethical and regulatory framework of the CLOZUK project, access to individual-level data requires a collaboration agreement with Cardiff University. Requests to access these datasets should be directed to Prof. James T. R. Walters.

## Acknowledgements

DBK and AFP were supported by an Academy of Medical Sciences “Springboard” award (SBF005\1083). The CLOZUK study was supported by the following grants from the Medical Research Council to Cardiff University: Centre (MR/L010305/1), Program (MR/P005748/1), and Project (MR/L011794/1, MC_PC_17212). It also received funding by the European Union’s Seventh Framework (279227, “CRESTAR”) and Horizon 2020 (964874, “REALMENT”) programs.

This work acknowledges the support of the Supercomputing Wales project, which is partly funded by the European Regional Development Fund (ERDF) via Welsh Government. It also acknowledges Lesley Bates, Catherine Bresner and Lucinda Hopkins (Cardiff University) for laboratory sample management, as well as Andy Walker (Magna Laboratories), Anoushka Colson (Leyden Delta) and Hreinn Stefansson (deCODE Genetics) for contributing to the sample collection, anonymization, data preparation and genotyping efforts of the CLOZUK2 sample.

## REFERENCES

1. Howes OD, McCutcheon R, Agid O, de Bartolomeis A, van Beveren Njm, Birnbaum ML, et al. (2017): Treatment-Resistant Schizophrenia: Treatment Response and Resistance in Psychosis (TRRIP) Working Group Consensus Guidelines on Diagnosis and Terminology. Am J Psychiatry 174: 216– 229.

2. Wagner E, Siafis S, Fernando P, Falkai P, Honer WG, Röh A, et al. (2021): Efficacy and safety of clozapine in psychotic disorders—a systematic quantitative meta-review. Transl Psychiatry 11: 487.

3. Legge SE, Hamshere M, Hayes RD, Downs J, O’Donovan MC, Owen MJ, et al. (2016): Reasons for discontinuing clozapine: A cohort study of patients commencing treatment. Schizophr Res 174: 113–119.

4. Nair B, MacCabe JH (2014): Making clozapine safer: current perspectives on improving its tolerability. Future Neurol 9: 313–322.

5. Whiskey E, Barnard A, Oloyede E, Dzahini O, Taylor DM, Shergill SS (2021): An evaluation of the variation and underuse of clozapine in the United Kingdom. Acta Psychiatr Scand 143: 339–347.

6. Siskind D, Siskind V, Kisely S (2017): Clozapine Response Rates among People with Treatment-Resistant Schizophrenia: Data from a Systematic Review and Meta-Analysis. Can J Psychiatry 62: 772–777.

7. Okhuijsen-Pfeifer C, Sterk AY, Horn IM, Terstappen J, Kahn RS, Luykx JJ (2020): Demographic and clinical features as predictors of clozapine response in patients with schizophrenia spectrum disorders: A systematic review and meta-analysis. Neurosci Biobehav Rev 111: 246–252.

8. Lauschke V, Ingelman-Sundberg M (2016): The Importance of Patient-Specific Factors for Hepatic Drug Response and Toxicity. Int J Mol Sci 17: 1714.

9. Stern S, Linker S, Vadodaria KC, Marchetto MC, Gage FH (2018): Prediction of response to drug therapy in psychiatric disorders. Open Biol 8: 180031.

10. Pardiñas AF, Owen MJ, Walters JTR (2021): Pharmacogenomics: A road ahead for precision medicine in psychiatry. Neuron. https://doi.org/10.1016/j.neuron.2021.09.011

11. Paternoster L, Tilling K, Davey Smith G (2017): Genetic epidemiology and Mendelian randomization for informing disease therapeutics: Conceptual and methodological challenges ((G. S. Barsh, editor)). PLOS Genet 13: e1006944.

12. Murray GK, Lin T, Austin J, McGrath JJ, Hickie IB, Wray NR (2021): Could Polygenic Risk Scores Be Useful in Psychiatry? JAMA Psychiatry 78: 210.

13. Richardson TG, Harrison S, Hemani G, Davey Smith G (2019): An atlas of polygenic risk score associations to highlight putative causal relationships across the human phenome. Elife 8. https://doi.org/10.7554/eLife.43657

14. Schizophrenia Working Group of the Psychiatric Genomics Consortium P, Ripke S, Walters JTR, O’Donovan MC (2020): Mapping genomic loci prioritises genes and implicates synaptic biology in schizophrenia. medRxiv 2020.09.12.20192922.

15. Schizophrenia Working Group of the Psychiatric Genomics Consortium P, Ripke S et al, Schizophrenia Working Group of the Psychiatric Genomics C (2014): Biological insights from 108 schizophrenia-associated genetic loci, 2014/07/25. Nature 511: 421–427.

16. Legge SE, Pardiñas AF, Walters JTR (2019): Genomic treatment response prediction in schizophrenia. Pers Psychiatry 413–422.

17. Pardiñas AF, Smart SE, Willcocks IR, Holmans PA, Dennison CA, Lynham AJ, et al. (2022): Interaction Testing and Polygenic Risk Scoring to Estimate the Association of Common Genetic Variants With Treatment Resistance in Schizophrenia. JAMA Psychiatry. https://doi.org/10.1001/jamapsychiatry.2021.3799

18. Smart SE, Kępińska AP, Murray RM, MacCabe JH (2021): Predictors of treatment resistant schizophrenia: a systematic review of prospective observational studies. Psychol Med 51: 44–53.

19. Schulte PFJ (2003): What is an Adequate Trial with Clozapine? Clin Pharmacokinet 42: 607–618.

20. Siskind D, Sharma M, Pawar M, Pearson E, Wagner E, Warren N, Kisely S (2021): Clozapine levels as a predictor for therapeutic response: A systematic review and meta-analysis. Acta Psychiatr Scand 144: 422–432.

21. Molden E (2021): Therapeutic drug monitoring of clozapine in adults with schizophrenia: a review of challenges and strategies. Expert Opin Drug Metab Toxicol 17: 1211–1221.

22. Porcelli S, Balzarro B, Serretti A (2012): Clozapine resistance: Augmentation strategies. Eur Neuropsychopharmacol 22: 165–182.

23. Pardiñas AF, Holmans P, Pocklington AJ, Escott-Price V, Ripke S, Carrera N, et al. (2018): Common schizophrenia alleles are enriched in mutation-intolerant genes and in regions under strong background selection. Nat Genet 50: 381–389.

24. Zheutlin AB, Dennis J, Linnér RK, Moscati A, Restrepo N, Straub P, et al. (2019): Penetrance and pleiotropy of polygenic risk scores for schizophrenia in 106,160 patients across four health care systems. Am J Psychiatry 176: 846–855.

25. Hamshere ML, Walters JTR, Smith R, Richards AL, Green E, Grozeva D, et al. (2013): Genome-wide significant associations in schizophrenia to ITIH3/4, CACNA1C and SDCCAG8, and extensive replication of associations reported by the Schizophrenia PGC. Mol Psychiatry 18: 708–712.

26. Pardiñas AF, Nalmpanti M, Pocklington AJ, Legge SE, Medway C, King A, et al. (2019): Pharmacogenomic variants and drug interactions identified through the genetic analysis of clozapine metabolism. Am J Psychiatry 176: 477–486.

27. Mayén-Lobo YG, Martínez-Magaña JJ, Pérez-Aldana BE, Ortega-Vázquez A, Genis-Mendoza AD, Dávila-Ortiz de Montellano DJ, et al. (2021): Integrative Genomic–Epigenomic Analysis of Clozapine-Treated Patients with Refractory Psychosis. Pharmaceuticals 14: 118.

28. Subramanian S, Völlm BA, Huband N (2017): Clozapine dose for schizophrenia. Cochrane Database Syst Rev 2017. https://doi.org/10.1002/14651858.CD009555.pub2

29. Couchman L, Morgan PE, Spencer EP, Flanagan RJ (2010): Plasma Clozapine, Norclozapine, and the Clozapine:Norclozapine Ratio in Relation to Prescribed Dose and Other Factors: Data From a Therapeutic Drug Monitoring Service, 1993–2007. Ther Drug Monit 32: 438–447.

30. Hubbard L, Lynham AJ, Knott S, Underwood JFG, Anney R, Bisson JI, et al. (2022): DRAGON-Data: A platform and protocol for integrating genomic and phenotypic data across large psychiatric cohorts. medRxiv 2022.01.18.22269463.

31. Chang CC, Chow CC, Tellier LCAM, Vattikuti S, Purcell SM, Lee JJ (2015): Second-generation PLINK: Rising to the challenge of larger and richer datasets. Gigascience 4. https://doi.org/10.1186/s13742-015-0047-8

32. Anderson CA, Pettersson FH, Clarke GM, Cardon LR, Morris AP, Zondervan KT (2010): Data quality control in genetic case-control association studies. Nat Protoc 5: 1564–1573.

33. Igo RP, Cooke Bailey JN, Romm J, Haines JL, Wiggs JL (2016): Quality Control for the Illumina HumanExome BeadChip. Curr Protoc Hum Genet 90. https://doi.org/10.1002/cphg.15

34. Graffelman J, Jain D, Weir B (2017): A genome-wide study of Hardy–Weinberg equilibrium with next generation sequence data. Hum Genet 136: 727–741.

35. Das S, Forer L, Schönherr S, Sidore C, Locke AE, Kwong A, et al. (2016): Next-generation genotype imputation service and methods. Nat Genet 48: 1284–1287.

36. Albitar O, Harun SN, Zainal H, Ibrahim B, Sheikh Ghadzi SM (2020): Population Pharmacokinetics of Clozapine: A Systematic Review. Biomed Res Int 2020: 1–10.

37. Yengo L, Sidorenko J, Kemper KE, Zheng Z, Wood AR, Weedon MN, et al. (2018): Meta-analysis of genome-wide association studies for height and body mass index in ~700000 individuals of European ancestry. Hum Mol Genet 27: 3641–3649.

38. Canela-Xandri O, Rawlik K, Tenesa A (2018): An atlas of genetic associations in UK Biobank. Nat Genet 50: 1593–1599.

39. Ge T, Chen C-Y, Ni Y, Feng Y-CA, Smoller JW (2019): Polygenic prediction via Bayesian regression and continuous shrinkage priors. Nat Commun 10: 1776.

40. Ruderfer DM, Charney AW, Readhead B, Kidd BA, Kähler AK, Kenny PJ, et al. (2016): Polygenic overlap between schizophrenia risk and antipsychotic response: a genomic medicine approach. The Lancet Psychiatry 3: 350–357.

41. Johnson PE (2019): rockchalk: Regression Estimation and Presentation. Retrieved from https://cran.r-project.org/package=rockchalk

42. Legge SE, Pardiñas AF, Helthuis M, Jansen JA, Jollie K, Knapper S, et al. (2019): A genome-wide association study in individuals of African ancestry reveals the importance of the Duffy-null genotype in the assessment of clozapine-related neutropenia. Mol Psychiatry 24: 328–337.

43. Conomos MP, Miller MB, Thornton TA (2015): Robust Inference of Population Structure for Ancestry Prediction and Correction of Stratification in the Presence of Relatedness. Genet Epidemiol 39: 276–293.

44. Robin X, Turck N, Hainard A, Tiberti N, Lisacek F, Sanchez J-C, Müller M (2011): pROC: an open-source package for R and S+ to analyze and compare ROC curves. BMC Bioinformatics 12: 77.

45. Hanley JA, McNeil BJ (1982): The meaning and use of the area under a receiver operating characteristic (ROC) curve. Radiology 143: 29–36.

46. Lewis CM, Vassos E (2020, May 18): Polygenic risk scores: From research tools to clinical instruments. Genome Medicine, vol. 12. BioMed Central Ltd., p 44.

47. de Leon J, Schoretsanitis G, Smith RL, Molden E, Solismaa A, Seppälä N, et al. (2021): An International Adult Guideline for Making Clozapine Titration Safer by Using Six Ancestry-Based Personalized Dosing Titrations, CRP, and Clozapine Levels. Pharmacopsychiatry. https://doi.org/10.1055/a-1625-6388

48. Bousman CA, Bengesser SA, Aitchison KJ, Amare AT, Aschauer H, Baune BT, et al. (2021): Review and Consensus on Pharmacogenomic Testing in Psychiatry. Pharmacopsychiatry 54: 5–17.

49. Li J, Yoshikawa A, Brennan MD, Ramsey TL, Meltzer HY (2018): Genetic predictors of antipsychotic response to lurasidone identified in a genome wide association study and by schizophrenia risk genes. Schizophr Res 192: 194–204.

50. Lisoway AJ, Chen CC, Zai CC, Tiwari AK, Kennedy JL (2021): Toward personalized medicine in schizophrenia: Genetics and epigenetics of antipsychotic treatment. Schizophr Res 232: 112–124.

51. Sun L, Pennells L, Kaptoge S, Nelson CP, Ritchie SC, Abraham G, et al. (2021): Polygenic risk scores in cardiovascular risk prediction: A cohort study and modelling analyses ((G. Hindy, editor)). PLOS Med 18: e1003498.

52. Ingelman-Sundberg M, Mkrtchian S, Zhou Y, Lauschke VM (2018): Integrating rare genetic variants into pharmacogenetic drug response predictions. Hum Genomics 12: 26.

53. Yanes T, McInerney-Leo AM, Law MH, Cummings S (2020): The emerging field of polygenic risk scores and perspective for use in clinical care. Hum Mol Genet 29: R165–R176.

