## Supplementary_Materials for "Genomic stratification of clozapine prescription patterns using schizophrenia polygenic scores"

Antonio F. Pardiñas

MRC Centre for Neuropsychiatric Genetics and Genomics, Division of Psychological Medicine and Clinical Neurosciences, Hadyn Ellis Building, Maindy Road, Cardiff University, Cardiff, UK, CF24 4HQ

SUPPLEMENTARY MATERIAL

Supplementary Tables 1-3

Supplementary Figures 1-3

Supplementary References

Supplementary Table 1. Descriptive statistics for demographic and clinical variables for each CLOZUK sample used.

| **CLOZUK2** | | | | **CLOZUK3** | | | |
|---|---|---|---|---|---|---|---|
| | | Mean | SD | | | Mean | SD |
| Age | | 42.98 | 11.10 | Age | | 42.97 | 11.08 |
| Highest clozapine dose (mg/day) | | 428.89 | 167.19 | Highest clozapine dose (mg/day) | | 429.47 | 167.22 |
| Clozapine concentration (ng/mL) | | 597.06 | 369.41 | Clozapine concentration (ng/mL) | | 597.29 | 369.96 |
| Frequency of assessments | | 3.56 | 3.53 | Frequency of assessments | | 3.34 | 2.66 |
| | | N | % | | | N | % |
| Sex | | | | Sex | | | |
| | Males | 2302 | 73.48 | | Males | 655 | 72.06 |
| | Females | 831 | 26.52 | | Females | 254 | 27.94 |
| Ancestry | | | | Ancestry | | | |
| | Admixed/Unknown | 84 | 2.68 | | Admixed/Unknown | 38 | 4.18 |
| | East Asian | 31 | 0.99 | | East Asian | 5 | 0.55 |
| | European | 2577 | 82.25 | | European | 761 | 83.72 |
| | North African | 92 | 2.94 | | North African | 26 | 2.86 |
| | Southwest Asian | 173 | 5.52 | | Southwest Asian | 42 | 4.62 |
| | Sub-Saharan African | 176 | 5.62 | | Sub-Saharan African | 37 | 4.07 |

Supplementary Table 2: Generalized linear regression model for highest clozapine daily dose adjusting for potential confounders in CLOZUK2

| | BETA | S.E. | P | $\Delta R^2$ |
|---|---|---|---|---|
| **PRS SCZ** | 11.348 | 3.597 | 0.0016 | 0.0027 |
| **AGE** | 8.973 | 1.626 | 3.71E-08 | 0.0084 |
| **$AGE^2$** | -0.103 | 0.018 | 1.19E-08 | 0.0090 |
| **FEMALE** | -57.016 | 6.383 | 7.00E-19 | 0.0219 |
| **PRS BMI** | 3.672 | 3.374 | 0.2766 | 0.0003 |
| **PRS SMOKING** | 5.032 | 2.829 | 0.0754 | 0.0009 |
| **PRS COFFEE** | 11.181 | 2.991 | 0.0002 | 0.0038 |
| **PC1** | 1.746 | 11.780 | 0.8822 | <0.0001 |
| **PC2** | 22.807 | 11.342 | 0.0444 | 0.0011 |
| **PC3** | -0.823 | 7.528 | 0.9130 | <0.0001 |
| **PC4** | 6.787 | 4.892 | 0.1655 | 0.0005 |
| **PC5** | 0.304 | 2.919 | 0.9172 | <0.0001 |
| **PC6** | 0.363 | 2.984 | 0.9033 | <0.0001 |
| **PC7** | 6.192 | 2.161 | 0.0042 | 0.0023 |
| **PC8** | -1.295 | 1.695 | 0.4449 | 0.0002 |
| **PC9** | -1.968 | 3.233 | 0.5428 | 0.0001 |
| **PC10** | 3.678 | 2.990 | 0.2187 | 0.0004 |
| **EAS** | 1.331 | 1.046 | 0.2035 | 0.0004 |
| **SAS** | -0.192 | 0.374 | 0.6070 | 0.0001 |
| **SSA** | -0.135 | 0.458 | 0.7679 | <0.0001 |
| **NEA** | 0.106 | 0.311 | 0.7333 | <0.0001 |
| **CLOZAPINE LEVELS** | 0.104 | 0.008 | 6.67E-41 | 0.0508 |
| **ASSESSMENTS** | 7.436 | 0.802 | 3.35E-20 | 0.0236 |

Supplementary Table 3: Generalized linear regression model for highest clozapine daily dose adjusting for potential confounders in CLOZUK3

| | BETA | S.E. | P | $\Delta R^2$ |
|---|---|---|---|---|
| **PRS SCZ** | 10.972 | 5.785 | 0.058 | 0.0035 |
| **AGE** | 5.899 | 2.400 | 0.014 | 0.0059 |
| **$AGE^2$** | -0.073 | 0.028 | 0.009 | 0.0068 |
| **FEMALE** | -30.315 | 10.989 | 0.006 | 0.0075 |
| **PRS BMI** | 10.610 | 7.507 | 0.158 | 0.0020 |
| **PRS SMOKING** | 5.222 | 4.931 | 0.290 | 0.0011 |
| **PRS COFFEE** | 14.439 | 5.073 | 0.005 | 0.0080 |
| **PC1** | -13.488 | 13.991 | 0.335 | 0.0009 |
| **PC2** | -9.828 | 17.536 | 0.575 | 0.0003 |
| **PC3** | 15.708 | 8.700 | 0.071 | 0.0032 |
| **PC4** | -5.526 | 15.133 | 0.715 | 0.0001 |
| **PC5** | -0.194 | 6.854 | 0.977 | <0.0001 |
| **PC6** | 14.061 | 5.732 | 0.014 | 0.0059 |
| **PC7** | -3.537 | 4.899 | 0.471 | 0.0005 |
| **PC8** | -10.104 | 5.078 | 0.047 | 0.0039 |
| **PC9** | -3.342 | 4.893 | 0.495 | 0.0005 |
| **PC10** | -5.942 | 4.646 | 0.201 | 0.0016 |
| **EAS** | 0.766 | 2.132 | 0.720 | 0.0001 |
| **SAS** | -0.110 | 0.870 | 0.899 | <0.0001 |
| **SSA** | 0.383 | 0.511 | 0.454 | 0.0006 |
| **NEA** | 0.591 | 0.616 | 0.337 | 0.0009 |
| **CLOZAPINE LEVELS** | 0.044 | 0.016 | 0.006 | 0.0074 |
| **ASSESSMENTS** | 6.609 | 0.899 | 4.37E-13 | 0.0531 |

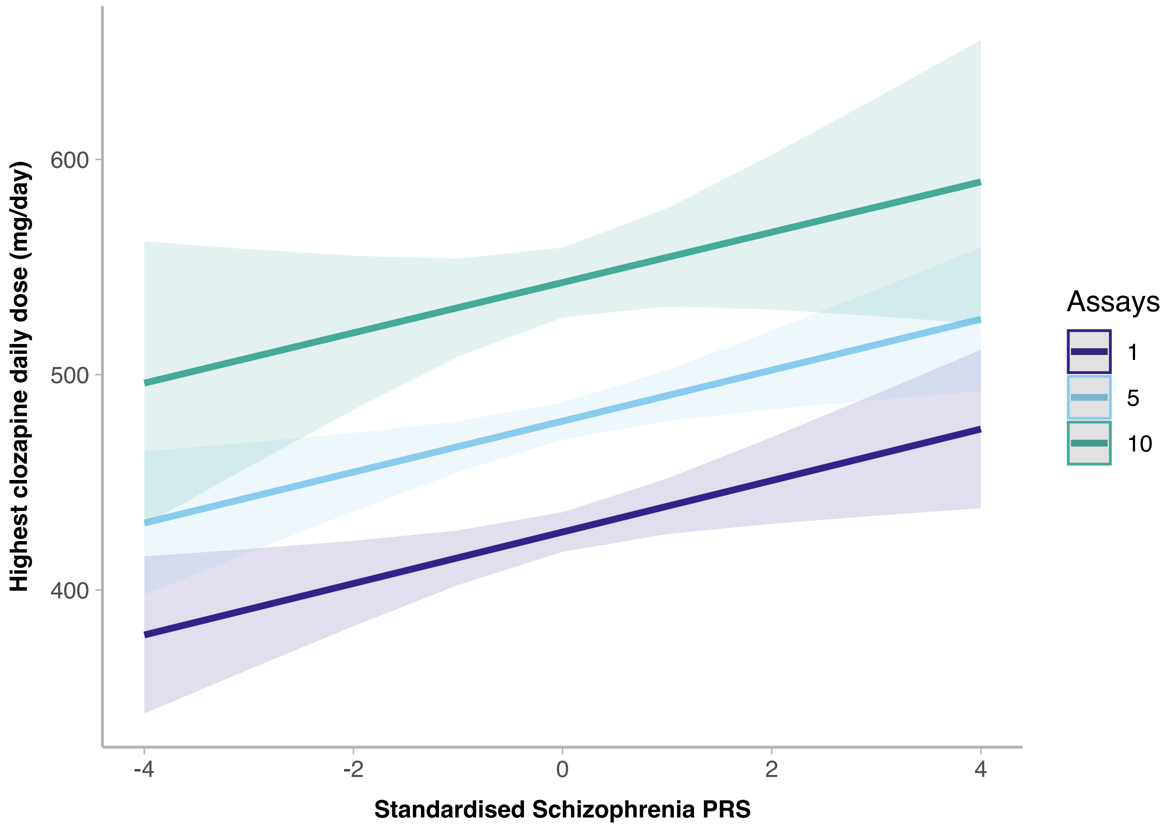


*Supplementary Figure 1 Effect of clozapine monitoring frequency on the relationship between schizophrenia PRS and highest clozapine daily dose*

*PRS = polygenic risk score*

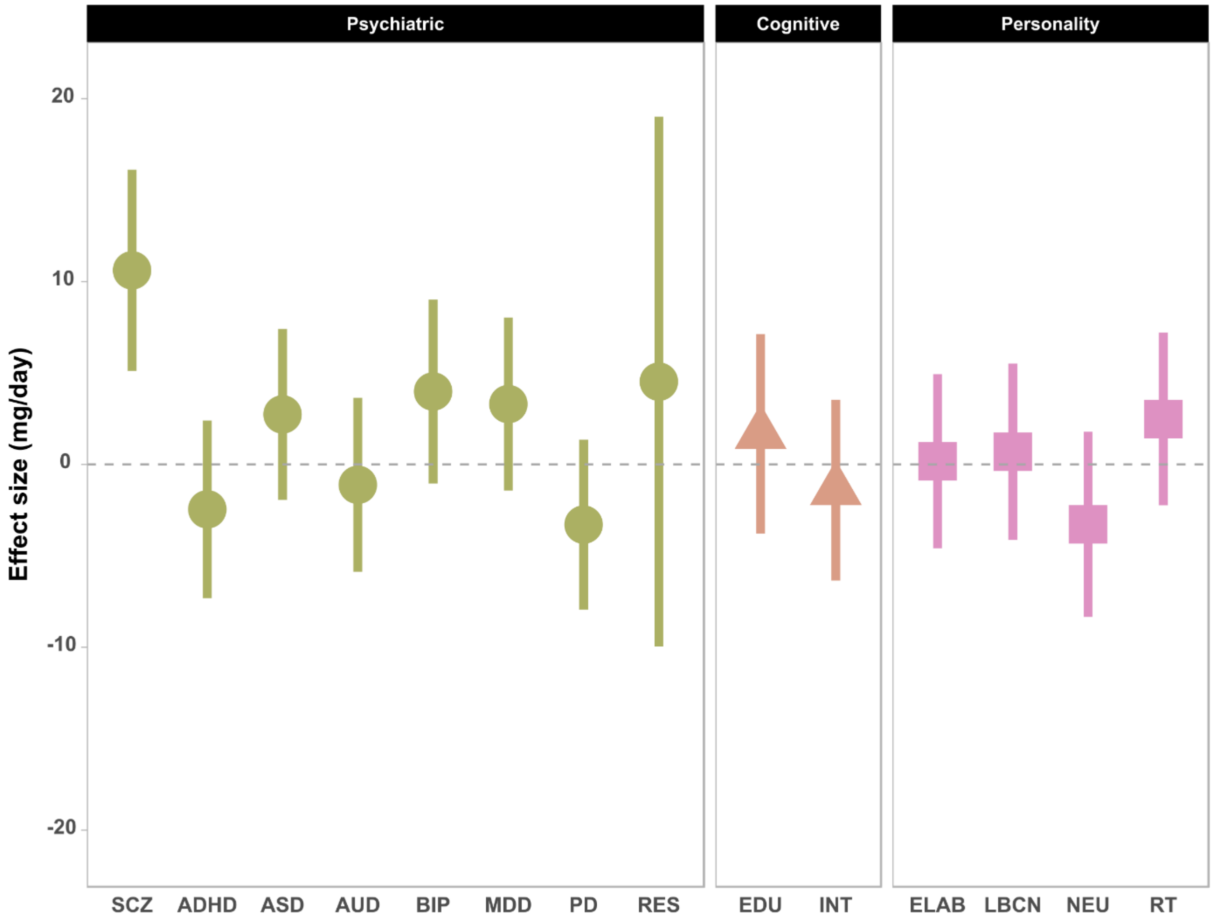


*Supplementary Figure 2 – Association of psychiatric, cognitive and personality PRS with highest clozapine doses. Underlying regressions use the same full model covariates as Supplementary Table 2 and Supplementary Table 3 and were computed on the merged CLOZUK2+3 sample for increased statistical power. All PRS were computed with the PRScs software (see methods).*

*SCZ: Schizophrenia (n= 130300)* (1)

*ADHD: Attention deficit hyperactivity disorder (n= 55374).*(2)

*ASD: Autism spectrum disorder (n= 46350).*(3)

*AUD: Alcohol use disorder (n= 121604).*(4)

*BIP: Bipolar disorder (n= 413466).*(5)

*MDD: Major depressive disorder (n= 807553).*(6)

*PD: Panic disorder (n= 10240).*(7)

*RES: Resilience to schizophrenia (n= 22405).*(8)

*EDU: Educational attainment (n= 765283).*(9)

*INT: Intelligence (n= 269867)*(10)

*ELAB: Emotional lability (n= 5133).*(11)

*LBCN: Lack of behavioral control (n= 5133).*(11)

*NEU: Neuroticism (n= 582989).*(12)

*RT: Risk-taking behavior (n= 939908).* (13)

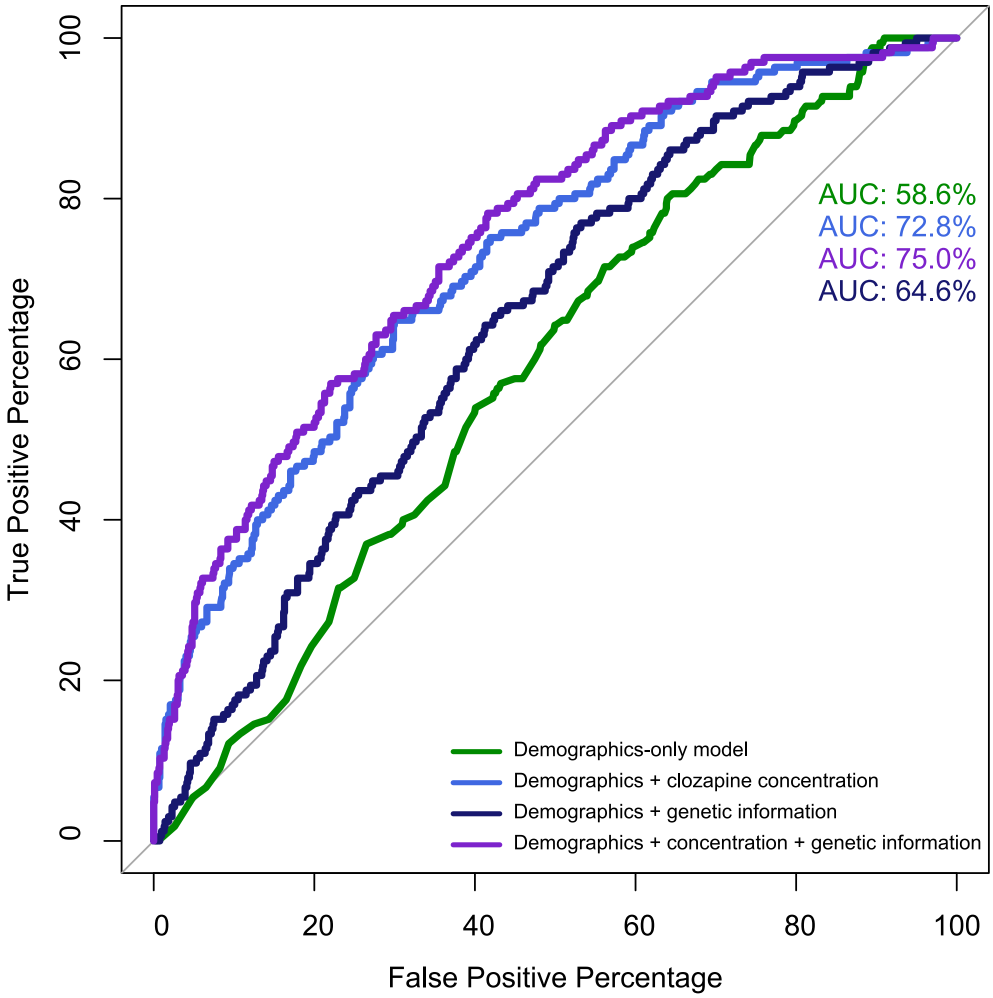


*Supplementary Figure 3 – ROC curves for prediction accuracy of probability of taking a high dose from models with different levels of information in CLOZUK2.*

*Demographics-only model included = sex, age, age2 and self-reported ethnicity*

*Demographics + clozapine included = sex, age, age2, self-reported ethnicity and clozapine plasma concentration*

*Demographics + genetics included = sex, age, age2, BMI PRS, Smoking PRS, Caffeine PRS, 10 genetic principal components and the probabilities of belonging to each biogeographic region (EAS, SAS, SSA, NEA)*

*Demographics+ clozapine concentration + genetic information included a full prediction model: sex, age, age2, BMI PRS, Smoking PRS, Caffeine PRS, 10 genetic principal components, the probabilities of belonging to each biogeographic region (EAS, SAS, SSA, NEA) and clozapine concentration levels*

### SUPPLEMENTARY MATERIAL REFERENCES


1. Schizophrenia Working Group of the Psychiatric Genomics Consortium P, Ripke S, Walters JTR, O'Donovan MC (2020): Mapping genomic loci prioritises genes and implicates synaptic biology in schizophrenia. *medRxiv* 2020.09.12.20192922.

2. Demontis D, Walters RK, Martin J, Mattheisen M, Als TD, Agerbo E, *et al.* (2019): Discovery of the first genome-wide significant risk loci for attention deficit/hyperactivity disorder. *Nat Genet* 51: 63–75.

3. Grove J, Ripke S, Als TD, Mattheisen M, Walters RK, Won H, *et al.* (2019): Identification of common genetic risk variants for autism spectrum disorder. *Nat Genet* 51: 431–444.

4. Sanchez-Roige S, Palmer AA, Fontanillas P, Elson SL, Adams MJ, Howard DM, *et al.* (2019): Genome-Wide Association Study Meta-Analysis of the Alcohol Use Disorders Identification Test (AUDIT) in Two Population-Based Cohorts. *Am J Psychiatry* 176: 107–118.

5. Mullins N, Forstner AJ, O'Connell KS, Coombes B, Coleman JRI, Qiao Z, *et al.* (2021): Genome-wide association study of more than 40,000 bipolar disorder cases provides new insights into the underlying biology. *Nat Genet* 53: 817–829.

6. Howard DM, Adams MJ, Clarke T-K, Hafferty JD, Gibson J, Shirali M, *et al.* (2019): Genome-wide meta-analysis of depression identifies 102 independent variants and highlights the importance of the prefrontal brain regions. *Nat Neurosci* 22: 343–352.

7. Forstner AJ, Awasthi S, Wolf C, Maron E, Erhardt A, Czamara D, *et al.* (2021): Genome-wide association study of panic disorder reveals genetic overlap with neuroticism and depression. *Mol Psychiatry* 26: 4179–4190.

8. Hess JL, Tylee DS, Mattheisen M, Børglum AD, Als TD, Grove J, *et al.* (2021): A polygenic resilience score moderates the genetic risk for schizophrenia. *Mol Psychiatry* 26: 800–815.

9. Okbay A, ..., Koellinger PD, ..., Turley P, Visscher PM, *et al.* (2022): Polygenic prediction within and between families from a 3-million-person GWAS of educational attainment. *Nat Genet* in press.

10. JE Savage PJSSKWJBCL (2018): Genome-wide association meta-analysis in 269,867 individuals identifies new genetic and functional links to intelligence. *Nat Genet* 50: 912–9.

11. Heilbronner U, Papiol S, Budde M, Andlauer TFM, Strohmaier J, Streit F, *et al.* (2021): "The Heidelberg Five" personality dimensions: Genome-wide associations, polygenic risk for neuroticism, and psychopathology 20 years after assessment. *Am J Med Genet Part B Neuropsychiatr Genet* 186: 77–89.

12. Baselmans BML, van de Weijer MP, Abdellaoui A, Vink JM, Hottenga JJ, Willemsen G, *et al.* (2019): A Genetic Investigation of the Well-Being Spectrum. *Behav Genet* 49: 286–297.

13. Karlsson Linnér R, Biroli P, Kong E, Meddens SFW, Wedow R, Fontana MA, *et al.* (2019): Genome-wide association analyses of risk tolerance and risky behaviors in over 1 million individuals identify hundreds of loci and shared genetic influences. *Nat Genet* 51: 245–257.
